# Reconstructing the course of the COVID-19 epidemic over 2020 for US states and counties: results of a Bayesian evidence synthesis model

**DOI:** 10.1101/2020.06.17.20133983

**Authors:** Melanie H. Chitwood, Marcus Russi, Kenneth Gunasekera, Joshua Havumaki, Fayette Klaassen, Virginia E. Pitzer, Joshua A. Salomon, Nicole A. Swartwood, Joshua L. Warren, Daniel M. Weinberger, Ted Cohen, Nicolas A. Menzies

## Abstract

Reported COVID-19 cases and deaths provide a delayed and incomplete picture of SARS-CoV-2 infections in the United States (US). Accurate estimates of both the timing and magnitude of infections are needed to characterize viral transmission dynamics and better understand COVID- 19 disease burden. We estimated time trends in SARS-CoV-2 transmission and other COVID-19 outcomes for every county in the US, from the first reported COVID-19 case in January 13, 2020 through January 1, 2021. To do so we employed a Bayesian modeling approach that explicitly accounts for reporting delays and variation in case ascertainment, and generates daily estimates of incident SARS-CoV-2 infections on the basis of reported COVID-19 cases and deaths. The model is freely available as the *covidestim* R package. Nationally, we estimated there had been 49 million symptomatic COVID-19 cases and 400,718 COVID-19 deaths by the end of 2020, and that 27% of the US population had been infected. The results also demonstrate wide county-level variability in the timing and magnitude of incidence, with local epidemiological trends differing substantially from state or regional averages, leading to large differences in the estimated proportion of the population infected by the end of 2020. Our estimates of true COVID-19 related deaths are consistent with independent estimates of excess mortality, and our estimated trends in cumulative incidence of SARS-CoV-2 infection are consistent with trends in seroprevalence estimates from available antibody testing studies. Reconstructing the underlying incidence of SARS-CoV-2 infections across US counties allows for a more granular understanding of disease trends and the potential impact of epidemiological drivers.

## Introduction

The numbers of newly diagnosed cases and confirmed COVID-19 deaths are the most easily observed measures of the health burden associated with COVID-19 and have been widely used to track the trajectory of the epidemic at the national, state, and local level.^1, 2^ However, there are at least three limitations of using reported cases and deaths for this purpose. First, testing is primarily organized to identify symptomatic individuals, but a large fraction of SARS-CoV-2 infections are asymptomatic,^3^ leading to case counts that are substantially smaller than the true incidence of infection. Second, the degree to which case counts undercount infections is sensitive to the availability and utilization of diagnostic testing, which has varied over time and geography.^4–6^ For this reason, it can be difficult to distinguish true trends from changes in testing practices. Third, case and death counts are lagging indicators of the transmission dynamics of the pathogen, as they are affected by delays associated with the incubation period, care-seeking behavior of symptomatic individuals, diagnostic processing times, and reporting practices. Taken together, these limitations present challenges to analyses that rely on these metrics as primary signals of SARS-CoV-2 spread.

A better indicator of changes in local transmission is the effective reproduction number (*R_t_*), which represents the average number of secondary infections caused by an individual infected at some time *t*.^7^ *R_t_* can signal short-term changes in transmission in response to policy and behavioral changes. However, *R_t_* is not a directly observable quantity and estimates of *R_t_* based on raw case reports become biased when reporting delays are incorrectly estimated,^5^ weakening their usefulness as a measure of transmission.

Unbiased estimates of COVID-19 cases and the *R_t_* of SARS-CoV-2 can provide more accurate insight into the size and scope of the United States (US) epidemic and inform current and future COVID-19 control policies. A number of modeling approaches have been developed to reconstruct the time series of infections and deaths over the course of the US epidemic. These approaches typically do not allow for variability in case ascertainment and infection fatality ratios (IFRs) across space and time, nor do they attempt to model SARS-CoV-2 infections or COVID-19 deaths at fine spatial scales, such as at the county level.^8, 9^

Here, we present detailed estimates of viral dynamics for all US states and counties, based on a Bayesian statistical model that combines multiple data sources to estimate SARS-CoV-2 infection patterns from observed case notifications and death reports. We apply our model to publicly available COVID-19 case and death data and report on the trajectory of the epidemic from the first reported case (January 13, 2020) until January 1, 2021. The model is available on GitHub (https://github.com/covidestim/covidestim/) as a package for the R programming language (*covidestim*).

## Results

### Analytic Overview

We developed a mechanistic model to back-calculate SARS-CoV-2 infections and subsequent outcomes based on reported COVID-19 cases and deaths. In this model the natural history of COVID-19 is represented using four health states: asymptomatic or pre-symptomatic SARS-CoV-2 infection (*Asymptomatic*), symptomatic but not severe COVID-19 disease (*Symptomatic*), severe COVID-19 disease (*Severe*), and death from COVID-19 (*Death*). In each health state (except *Death*) individuals either recover or transition to a more severe state after some delay.

Infected individuals can be diagnosed in the *Asymptomatic*, *Symptomatic*, or *Severe* states, and we assume all diagnosed cases and all deaths among diagnosed individuals are reported after as short delay. Figure 1 shows modeled health states and transitions. The model generates several outcomes of epidemiological importance, including *R_t_*, total infections, symptomatic cases, total deaths, and case ascertainment; we estimated these outcomes for each US state and county from the start of the epidemic until January 1, 2021.

**Figure 1:**
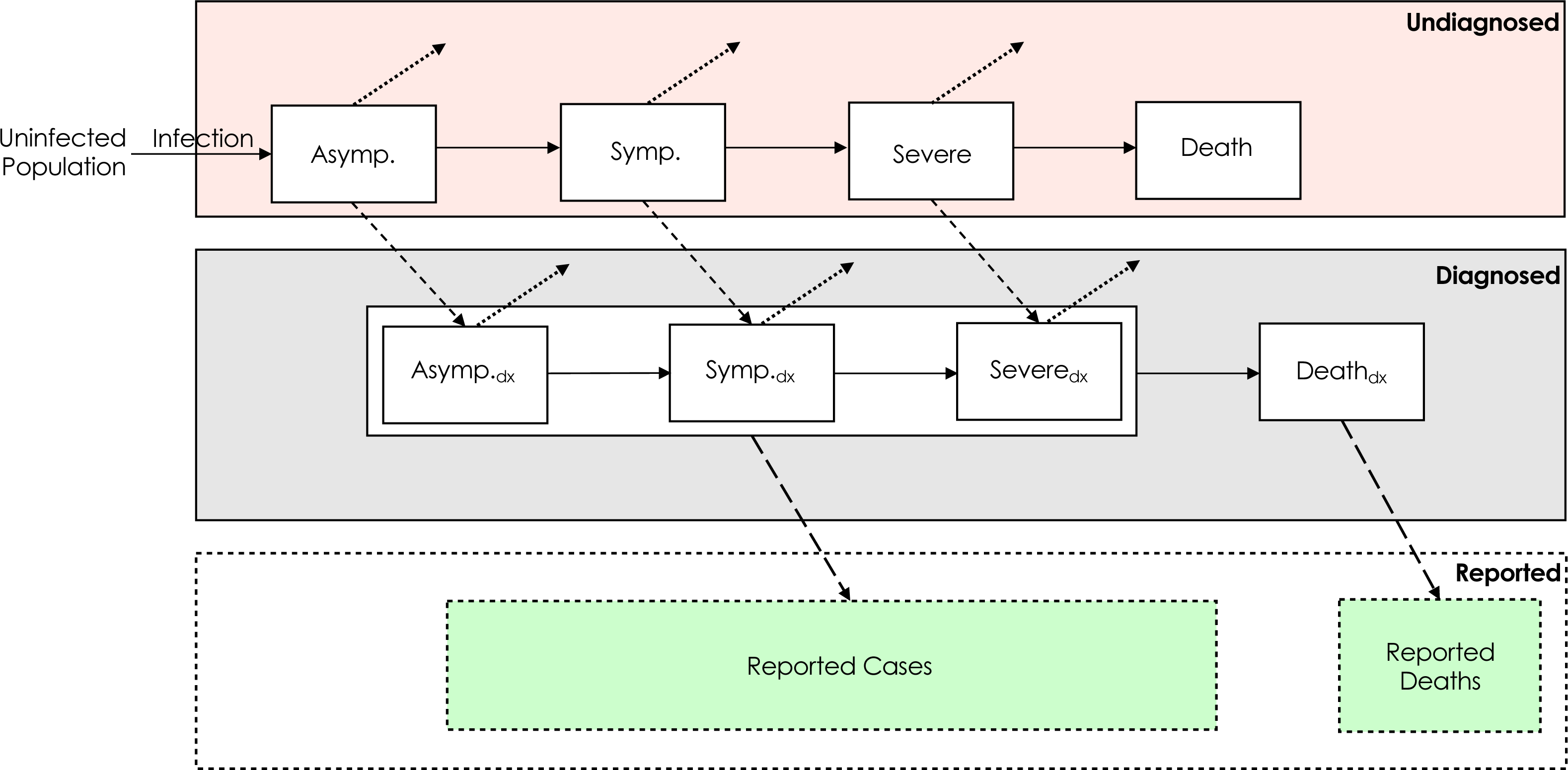
A model schematic of the main health states: *Asymptomatic* (denoted “Asymp.”), *Symptomatic* (denoted “Symp.”), *Severe*, and *Death*. The subscript “dx” indicates that individuals in that state have received a diagnosis of COVID-19. Each transition (denoted with an arrow) has an associated probability and delay distribution. Solid arrows denote disease progression; dotted arrows denote recovery; short dashed arrows denote diagnosis; long dashed arrows denote reporting. All diagnosed cases and deaths are assumed to be reported after a given delay.

### Main Findings

#### Incidence and R_t_

The SARS-CoV-2 epidemic in the US consisted of a series of related outbreaks, which varied greatly in both the intensity of transmission and the extent of geographic spread (Figure 2). The March outbreak in New York State was the largest per population in a single state; on March 28, we estimate that New York State had 812 (95% credible interval: 477, 1393) infections per 100,000, and 39% (23%, 68%) of all infections in the US on that day. Local surges in infections during the fall and winter of 2020 rivaled New York’s March outbreak in scale, but occurred in the context of a more generalized US epidemic. South Dakota, for example, had its highest per capita infections of 2020 on November 7 (628 [405, 1009] infections per 100,000), but accounted for just 1.2% (0.8%, 2.0%) of all US infections that day. Forty-five states experienced the highest daily infections per capita in November or December (Figure 3).

**Figure 2:**
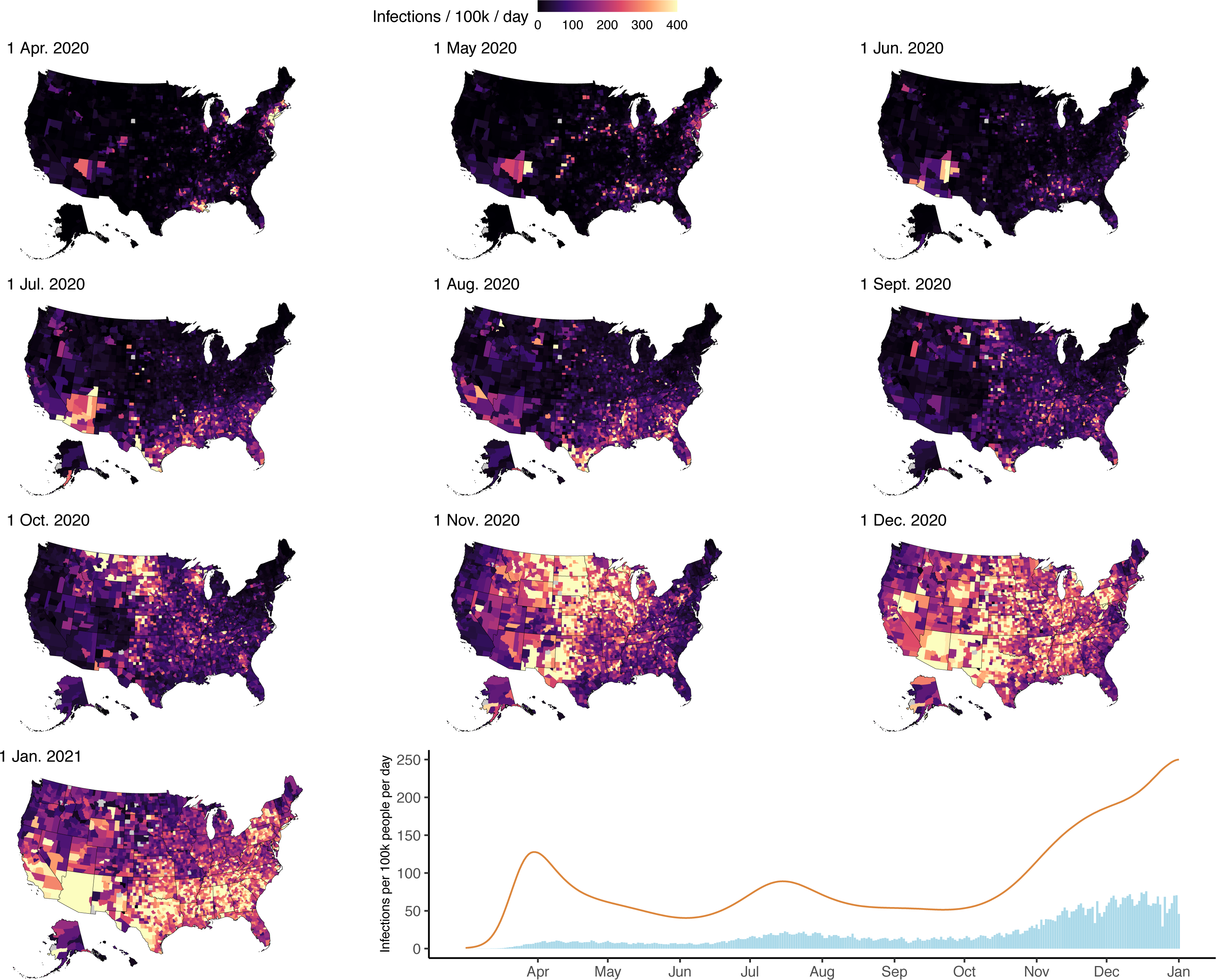
Panels 1-10: County-level infections per 100,000 population per day at 10 timepoints between April 1, 2020 and January 1, 2021. Panel 11: Time series of national SARS-CoV-2 infection estimates (orange line) and reported COVID-19 diagnoses (blue bars) per 100,000 people per day from March 1, 2020 to January 1, 2021.

**Figure 3:**
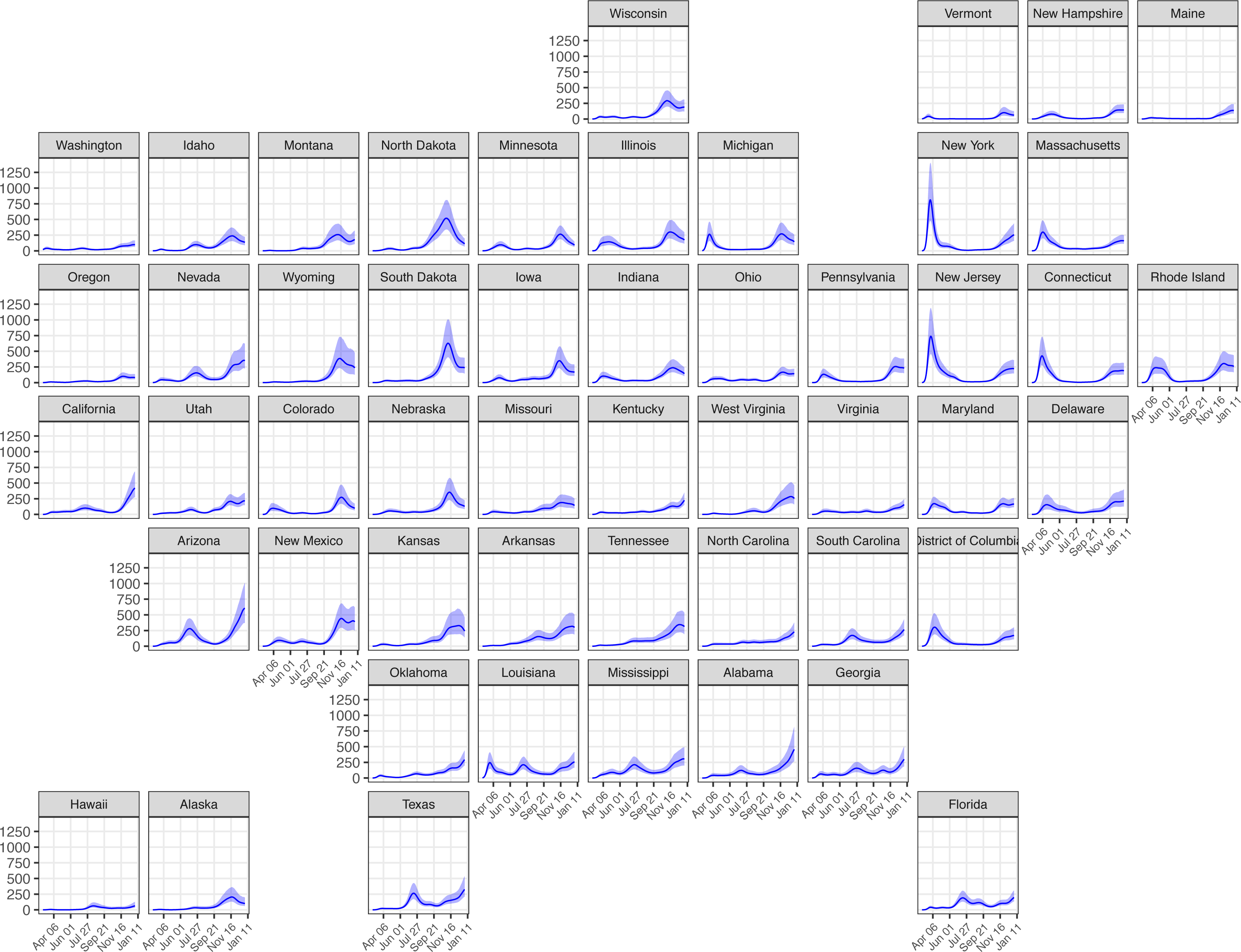
Incident infections per 100,000 residents per day for each US state from March 1, 2020 to January 1, 2021. Shaded areas represent 95% credible intervals.

While most states and counties had lower levels of transmission during the summer months, few achieved established thresholds of low levels of community transmission, defined as fewer than 20 confirmed cases per 100,000 per week.^10^ We estimate that only six states (Alaska, Hawaii, Montana, Oregon, Vermont, and West Virginia) had fewer than 20 symptomatic cases per 100,000 inhabitants per week after transmission was established locally. Notably, Vermont remained below this threshold from the week of April 27 until the week of October 5.

Estimates of *R_t_* at the start of the epidemic varied greatly by state. The median state-level estimate of *R_t_* on the first day a case was reported in each state was 4.9 (range: 2.0 [1.7 – 2.4] in Washington to 11.4 [7.4– 18.9] in New York). Throughout April, *R_t_* estimates dropped substantially. Over the period May 1, 2020 to January 1, 2021, state-level estimates of *R_t_* ranged from 0.6 (0.5, 0.8) to 1.7 (1.4, 2.0) (Figure 4).

**Figure 4:**
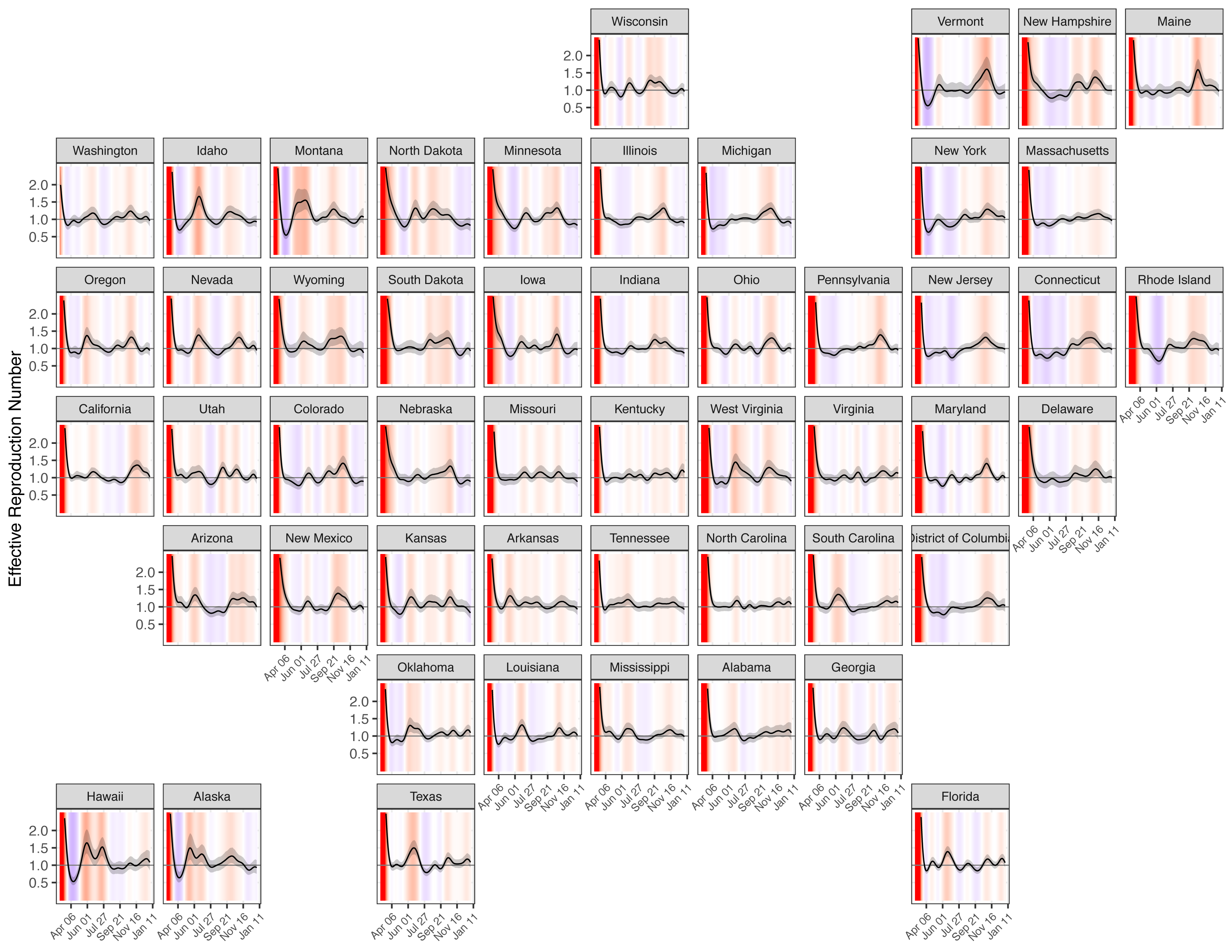
*R_t_* estimates for each US state from March 1, 2020 to January 1, 2021. Background colors indicate whether *R_t_* is substantially greater than 1 (red), close to 1 (white), or substantially less than 1 (blue). Grey line indicates *R_t_* = 1. Shaded areas represent 95% credible intervals.

#### Percent Ever-Infected with SARS-CoV-2

For each county, we calculated the percentage of the population ever-infected as the sum of all estimated infections divided by county population on January 1, 2021 (Figure 5). This cumulative infection estimate is distinct from reported seroprevalence estimates, as seroprevalence measures may be affected by the lower immune response among individuals with mild/asymptomatic infection, possible waning of antibody titers,^11, 12^ and non-representativeness of sampled populations.^13^ By January 1 2021, we found that the percent of the population ever-infected exceeded 50% in 241 (7.7%) counties and exceeded two-thirds of the population in 22 (0.7%) counties. Conversely, the percent ever-infected was less than 10% in 145 (4.6%) counties and less than 5% in 34 (1.1%) counties. Based on the sum of state estimates (posterior medians), we estimate that 27% of the US population had been infected with SARS-CoV-2 by January 1, 2021. Across states, the percent ever-infected ranged from 6.7% (4.4%, 10.9%) in Vermont to in 44.6% (30.0%, 66.5%) Arizona (Figure 3).

**Figure 5:**
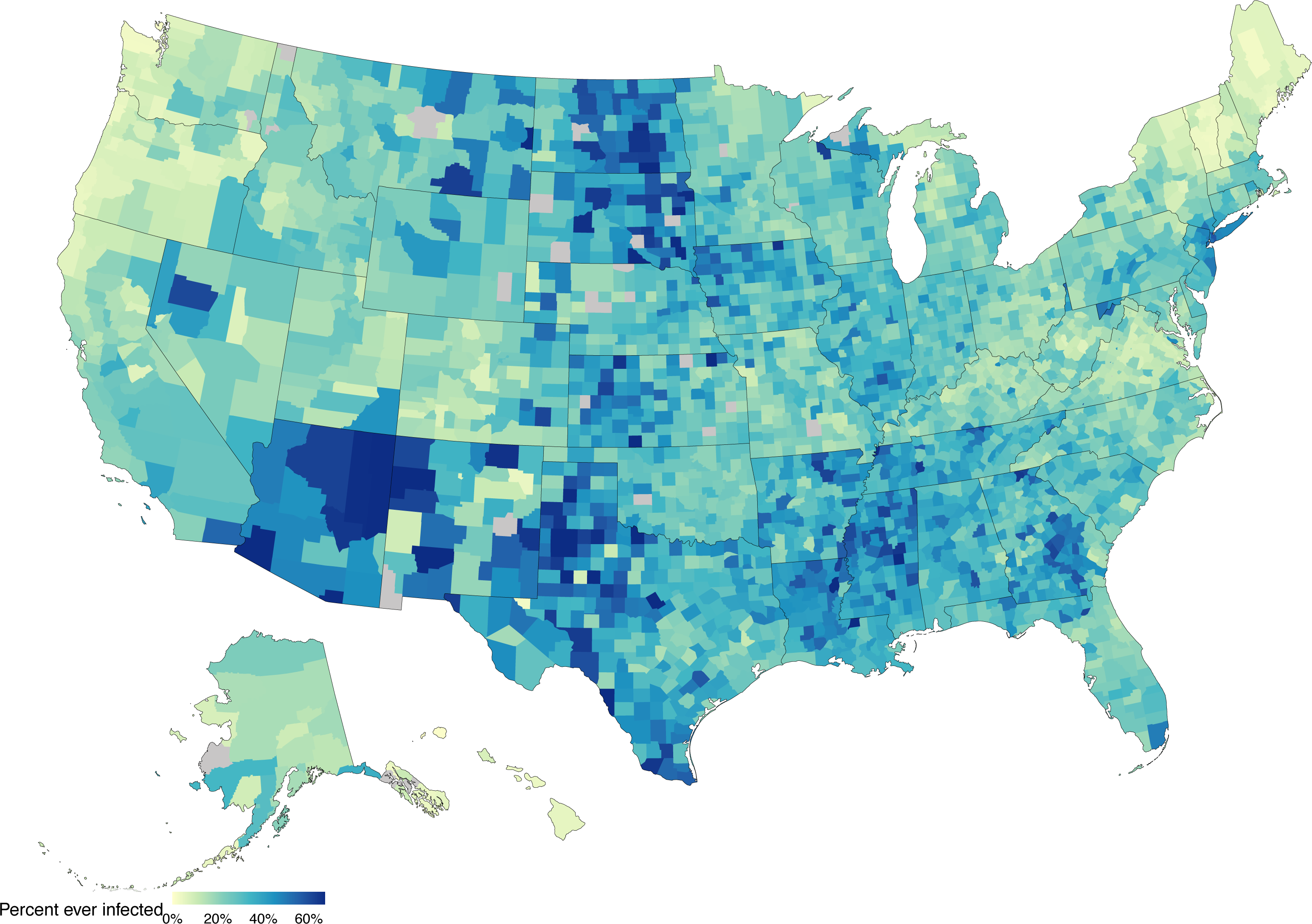
Percentage of the population ever-infected with SARS-CoV-2 as of January 1, 2021.

On January 1, 2021, the US had reported 349,247 cumulative COVID-19 deaths.^14^ Based on the sum of state estimates (posterior medians), we estimate there were 400,718 cumulative COVID- 19 deaths as of January 1, 2021, 14.7% greater than cumulative reported deaths and approximately 0.12% of the US population on January 1, 2020. Estimates of the size of the infected population were sensitive to assumptions about the infection fatality rate, with higher IFR values producing lower estimates of the infected population (Figure S1).

#### Case Ascertainment

The probability that an infection is diagnosed changed substantially over the course of the U.S. epidemic. Ascertainment was low in the months of March, April, and May 2020. The national median state-level case ascertainment (based on state-level posterior medians) in this period was 14% (range: 3.8%, 41.2%). Infection ascertainment improved steadily through June 2020 before plateauing; the national mean probability of diagnosis fluctuated between 24% and 37% between July 1, 2020 and January 1, 2021. Infection ascertainment estimates varied significantly across states, and state-level estimates were highly uncertain (Figure 6). Only 5 states achieved greater than 50% ascertainment at any point in time (based on posterior median). State-level model estimates of infection ascertainment each day are very weakly correlated with the seven-day moving average fraction of tests that have a positive result (Spearman rank correlation ( ) = - 0.10, p < 0.001). From the introduction of SARS-CoV-2 in the US until January 1, 2021, we estimate that 22.9% of infections were identified and reported.

**Figure 6:**
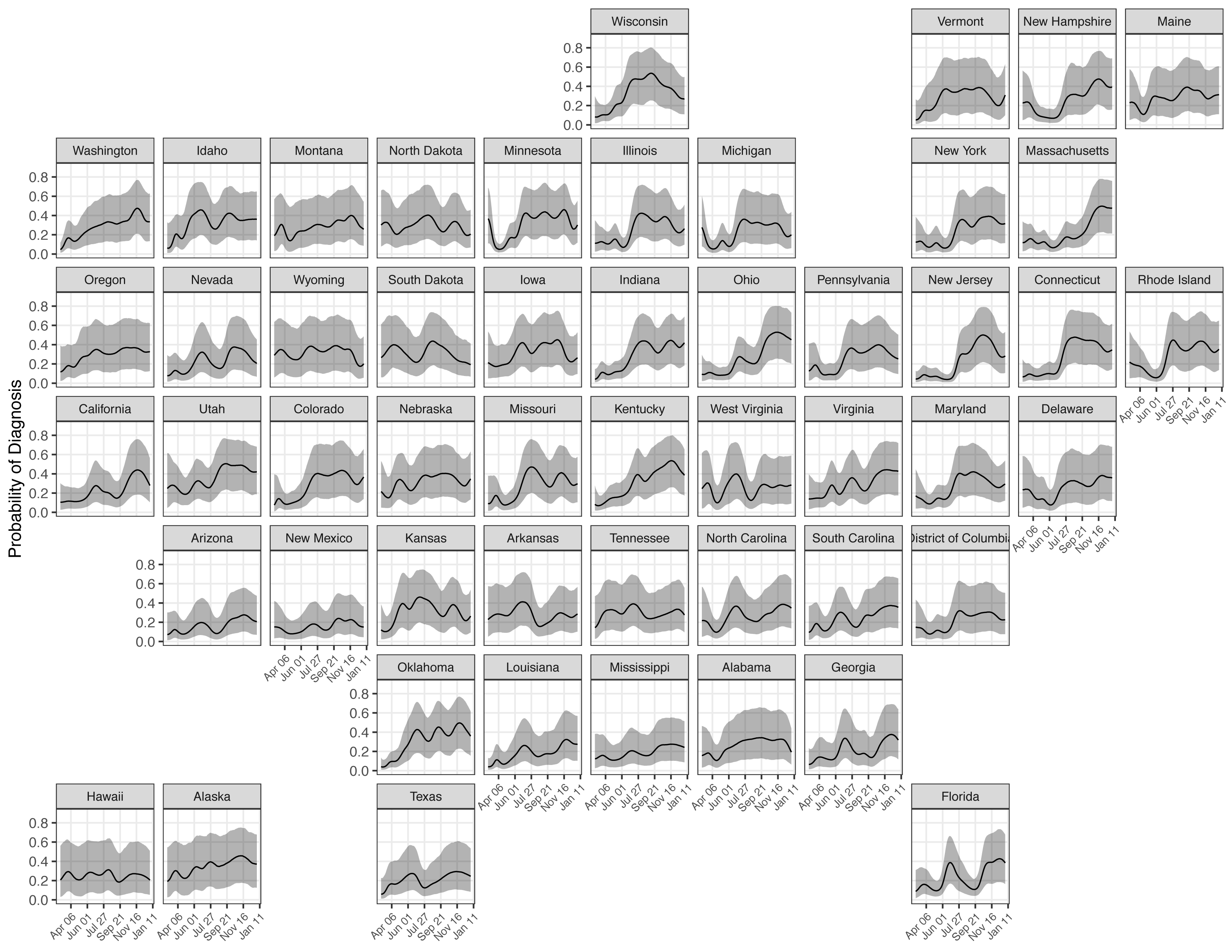
The probability that a person infected with SARS-CoV-2 on a given day will be diagnosed for each US state from March 1, 2020 to January 1, 2021. Shaded areas represent 95% credible intervals.

#### Comparisons to External Covid-19 Burden Indicators

We compared our estimates of the percent ever-infected with SARS-CoV-2 to U.S. Centers for Disease Control (CDC) seroprevalence estimates drawn from commercial laboratory data,^15^ acknowledging previously noted differences between these different outcomes. Derived from a convenience sample of blood specimens collected for reasons unrelated to COVID-19, the seroprevalence estimates provide state-level evidence on SARS-CoV-2 antibody test positivity at multiple time points (Figure 7). However, these estimates are incomplete in some states (e.g. South Dakota), and the series of values declines over time in others (e.g. New York). Comparing these estimates to other reported indicators of cumulative disease burden on January 1 2021, the modeled estimates of the percent ever-infected were more strongly correlated with cumulative hospitalizations (Spearman rank correlation (ρ) = 0.60) and cumulative reported deaths (ρ = 0.81) than the CDC seroprevalence estimates (ρ = 0.41 and 0.37 for hospitalizations and deaths respectively).

**Figure 7:**
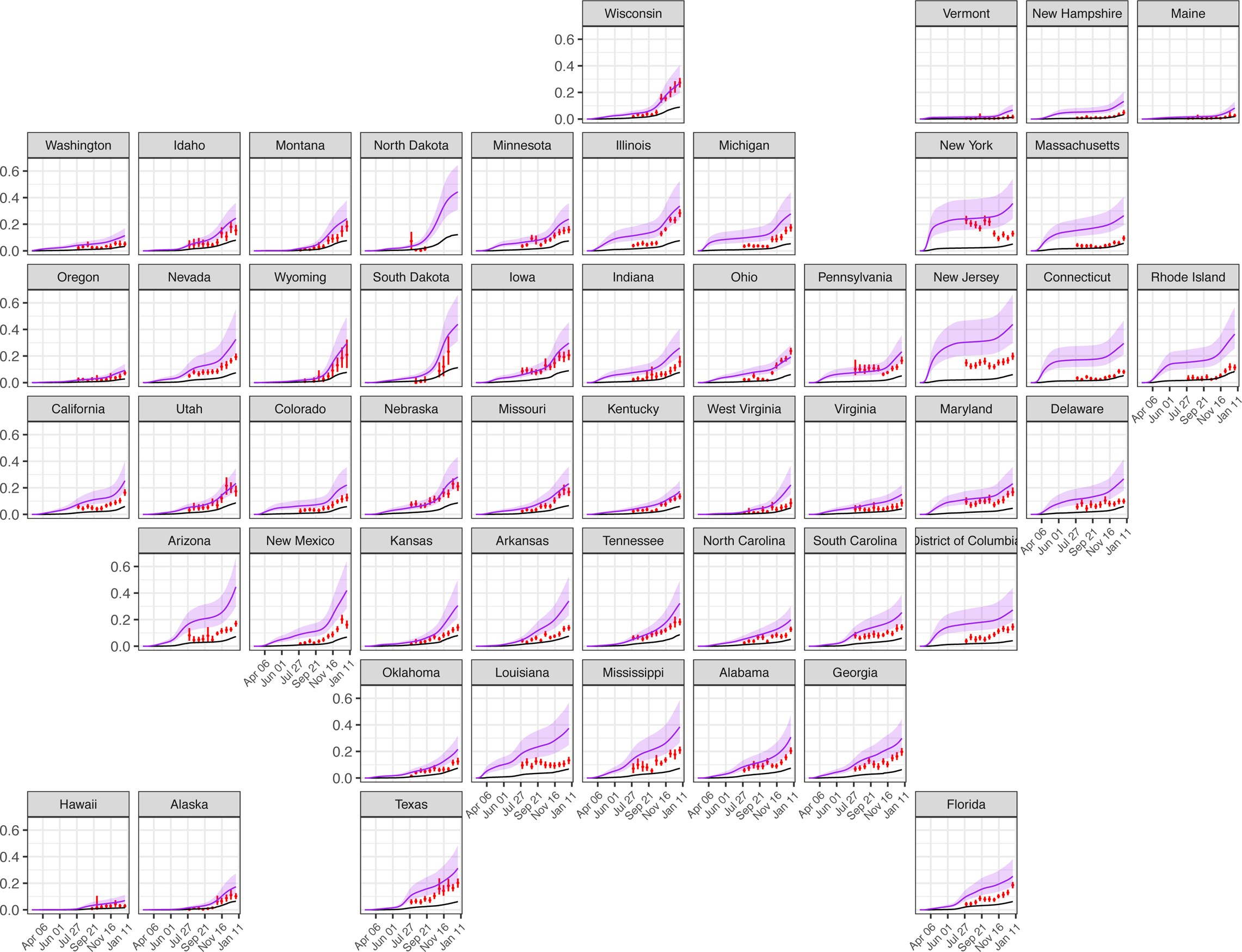
Comparison of the estimated percent ever-infected with SARS-CoV-2 (purple line, shaded areas represent 95% credible intervals) to CDC seroprevalence estimates from commercial laboratory data (red vertical line) and cumulative reported cases (black line) for each US state from March 1, 2020 to January 1, 2021.

In addition, we compared model estimates of cumulative COVID-19 deaths (detected and undetected) to state-level estimates of excess all-cause mortality, which reflect both COVID-19 deaths and deviations from expected levels and patterns in non-COVID-19 deaths,^6^ (Figure 8) at each weekly timepoint from March 7 to December 19, 2020. On average, modeled estimates of cumulative COVID-19 deaths are less than or approximately equal to estimates of excess all-cause mortality. Notably, three states (Alaska, Hawaii, Maine) have extended periods where the estimated all-cause mortality did not exceed all-cause mortality from previous years (i.e. excess mortality was negative); in periods where all-cause mortality is higher than expected, our estimates of COVID-19 deaths correlate strongly with excess mortality estimates (Spearman rank correlation (ρ) = 0.96, p < 0.001). Additionally, model estimates of cumulative COVID-19 deaths exceed estimates of excess all-cause mortality in two states (Massachusetts and Rhode Island). Estimates of excess all-cause mortality were not available for Connecticut, North Carolina, or West Virginia.

**Figure 8:**
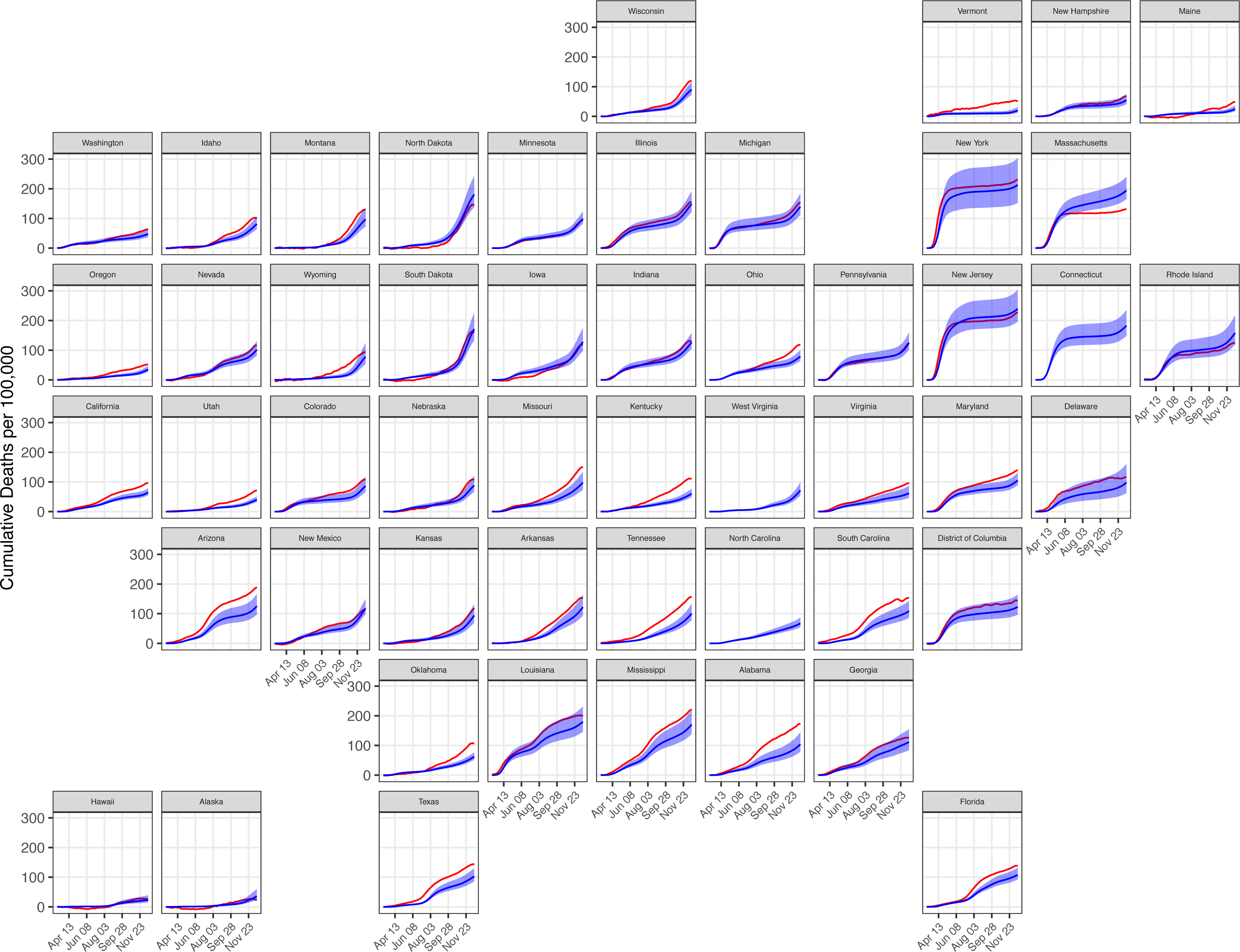
Comparison of cumulative COVID-19 deaths (blue) to cumulative excess all-cause mortality (red) for each US state from March 7 to December 19, 2020. Shaded areas represent 95% credible intervals.

## Discussion

We present detailed estimates of the dynamics of SARS-CoV-2 infections in US states and counties through the end of 2020. We found that the viral dynamics are best described as a series of related local and regional epidemics, differing in their timing and magnitude even within individual states. This is evident in the large variation in state- and county-level estimates of percent ever-infected as of January 1, 2021. As case ascertainment has also varied over space and time, these estimates provide insights beyond those that can be inferred from cumulative case counts alone. Ascertainment of infection improved markedly after the first months of the US epidemic, but remained low nationally; we conclude that the reported cumulative case count was approximately one-quarter of the true number of US infections at the end of 2020.

Most notably, we found that model estimates of cumulative infections differ from seroprevalence estimates produce by the CDC. We note that our estimates of cumulative infections are more strongly correlated with cumulative hospitalizations and deaths across states, potentially reflecting biases in the empirical seroprevalence estimates. Seroprevalence studies have a number of known limitations, including the use of non-representative samples^13^ and possible reduced sensitivity associated with waning of antibody titers, as has been reported for some tests.^11, 12^ A comparison between model estimates and seroprevalence data therefore suggests that this method provides valuable information about the incidence of infection over time.

The Bayesian estimation approach used for this analysis makes a number of simplifying assumptions. To reduce model complexity, we rely on fixed distributions to describe delays in disease progression and detection. Because we anchor the analysis on death data (under the assumption that deaths were more consistently reported than cases over the course of the epidemic), model estimates are sensitive to infection fatality risk (IFR) estimates, which are themselves uncertain. Finally, we assume that a previously infected individual cannot be re- infected with SARS-CoV-2. While waning antibody titers suggest that re-infection is possible over time, we do not believe that our assumptions about re-infection meaningfully impact our results.^16, 17^

In addition, we used data that have been aggregated from state-level reporting mechanisms, which are vulnerable to a number of potential sources of bias. States vary in their reporting criteria (e.g. reporting the number of positive tests as opposed to number of individuals who have tested positive) and the average delay between case detection and reporting. Data are also subject to occasional revisions, often implemented as a single-day change in the cumulative count of cases or deaths. Taken together, these data irregularities lead to additional variance in the reported data and a reduction in the precision of reported estimates. While line-list data would likely improve the precision of model estimate^18^, these data are not widely available in the US, and it is not possible to generate the results presented here from a method which relies solely on line-list data. Despite these limitations, the method described here may represent an improvement over similar modeling approaches that do not allow for case ascertainment rates and infection fatality ratios that vary over both space and time,^8, 9, 19, 20^ or that estimate *R_t_* using model outputs rather than as part of the modeling framework.^9, 18, 20^ Furthermore, our approach uses changes in case and death data to estimate changes in transmission, while others approaches make use of more indirect data on mobility^8, 19^ or similar proxies^20^ to signal changes in transmission. While mobility has a mechanistic relationship with disease transmission, the association between movement data and viral transmission is complex, possibly because of changes in mask use and other non-pharmaceutical interventions, and was an increasingly poor predictor of transmission as the epidemic progressed in the US.^21, 22^

In conclusion, the modeling approach described here provides a coherent framework for simultaneously estimating the trend in SARS-CoV-2 infections and the fraction of the population that has been infected previously, providing key information on the viral dynamics at county- and state-levels. While the deployment of effective vaccines against the virus represents a great hope for the control of SARS-CoV-2 transmission, vaccine hesitancy and the emergence of more transmissible variants^23^ present an ongoing challenge to disease control in the US. Understanding the course of the epidemic in the pre-vaccine era can help guide decision making in a landscape with heterogenous vaccine coverage. Ongoing, local evidence on trends in *R_t_* and new and cumulative infections will continue to be important for both governments and individuals.

## Methods

We developed a mechanistic model that uses reported case and death data to back-calculate the natural history cascade of SARS-CoV-2. The model estimates the expected number of cases and deaths reported on a given day as the convolution of the time series of diagnosed cases and deaths (among diagnosed individuals) and fixed reporting delay distributions; the expected number of diagnoses on a given day is estimated with health-state specific and time-varying probabilities of diagnosis. The model represents the natural history of COVID-19 as a series of health state transitions with associated probabilities and delays (Figure 1). The model utilizes delay distributions associated with health state progression, time-invariant probabilities of transitioning from *Asymptomatic* to *Symptomatic* and from *Symptomatic* to *Severe*, and a time-varying probability of transitioning from *Severe* to *Death*. The number of individuals entering *Asymptomatic* is a function of the serial interval, the fraction of the population not yet infected, and *R_t_*; *R_t_* is modeled using a log-transformed cubic b-spline.

### Data

For every state and county in the United States, we extracted daily data on reported COVID-19 cases and deaths from a repository compiled by the Johns Hopkins Center for Systems Science and Engineering (CSSE)^9^. We calculated the time series of new cases and deaths as the difference between cumulative counts reported on consecutive days. In instances in which the reported cumulative count decreased from one day to the next, we assumed that there were zero new cases or deaths on each day until the cumulative count exceeded the previous maximum. In several instances the data reported by CSSE fail to capture the beginning of the epidemic in early 2020, or exhibit irregularities during this period. To reconstruct the time series for this period we used data compiled by the Covid Tracking Project.^24^

### Mathematical model

We constructed a deterministic mathematical model relating reported cases and deaths to unobserved COVID-19 natural history. A flexible function for *R_t_* determines the number of individuals infected on a given day, and the model then tracks the progression of the infected cohort through health states of increasing disease severity, with modelled quantities—*A_t_* (*Asymptomatic*), *S_t_* (*Symptomatic*), *V_t_* (*Severe*), and *D_t_* (*Death*)—reflecting the number of individuals entering a given health state on day *t*. From each health state, an individual can either recover or progress to the next health state, with this transition governed by a defined delay distribution. Ultimately, the model estimates an expected number of reported cases and deaths on each day, which are fit to observed data via negative binomial likelihood functions.

#### New infections

We modelled the daily number of newly-infected individuals (*A_t_*) entering the *Asymptomatic* state. For each modelled location, we specified a random intercept (*A*_0_) 28 days before the first reported COVID-19 case, and calculated changes in as a function of the effective reproduction number (*R_t_* ) and the serial interval (*z*), measured in days.

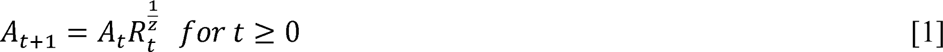

We modelled the time trend in *R_t_* using a log-transformed cubic b-spline (*X_RT_*) with knots every 5 days, allowing flexibility in the evolution of the epidemic curve over time. Penalties on first and second differences of the spline parameters were used to dampen oscillations not supported by the data. We assumed that individuals can only be infected once and multiplied the spline by the fraction of the population (*N*) uninfected at each timepoint, penalizing *R_t_* towards zero as the population ever-infected approaches 100%.

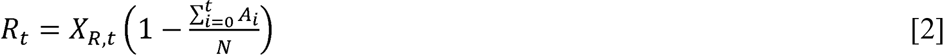

#### Disease progression

We assumed that a fraction of individuals with asymptomatic disease (*ps*) progress to the *Symptomatic* state. The delay from infection to symptoms was assumed to follow a Gamma distribution, with *psi*, representing the fraction progressing between *i* and *i+1* days after infection, among those progressing to the symptomatic state.

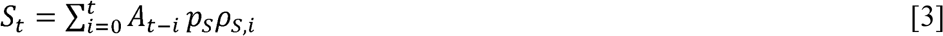

Similarly, a fraction of individuals in the *Symptomatic*state (*pv*) were assumed to progress to the *Severe* state, with Gamma-distributed delay distribution ,*ps,i* . A fraction of individuals with severe disease (*p_Dt_* ) die, with Gamma-distributed delay distribution

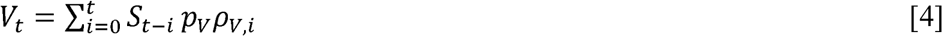

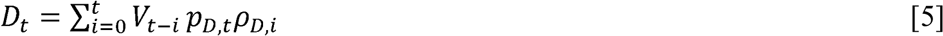

With the exception of *p_Dt_*, disease progression parameters were not allowed to vary over time. For *p_Dt_*, we assumed higher values applied in early 2020, reflecting higher case fatality among individuals with severe disease early in the epidemic due to later presentation and lower effectiveness of treatment at that time. We modeled the time trend in *p_Dt_*, as a sigmoid curve (operationalized using the Normal cumulative distribution function Φ) with an inflection point on May 1 2020. The odds ratio *OR_pd_*_b_ describes the elevated case fatality early in the epidemic, and *pd0*defines the progression probability after early 2020. In Equation 6, *μ* is equal to the number of days between the start of the model (*t*=0) and May 1^st^ 2020, and σ is equal to 21 days.

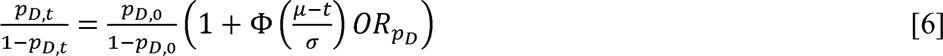

While vaccination would also affect disease progression probabilities, we assumed that vaccination coverage was insufficient to impact disease natural history during the study period.

### Infection fatality ratio

We assumed that the infection fatality ratio (IFR) differs across states and counties, reflecting differences in the age distribution of the epidemic and differences in the prevalence of medical risk factors for severe COVID-19 disease. First, we calculated the age distribution of infections for each state, based on the reported age distribution of COVID-19 deaths^25^ and published age-specific IFRs.^26^ Second, we used these age distributions to calculate an average IFR for each state, weighting the age-specific IFRs by the fraction of the population in each age group. This produced a national average IFR of 0.35, which we believe to be implausibly low; we rescaled state-level values to produce a national average IFR of 0.5%.^27^ As the age-distribution of COVID-19 deaths was not available at the county-level, we estimated county-level IFR values by multiplying the state-average IFR by the prevalence of medical risk factors for severe COVID-19 disease in each county relative to the rest of the state.^28^ To understand the implications uncertainty in the IFR for modelled estimates of the infected population, we plotted the relationship between these two quantities in the fitted model outcomes.

### Diagnosis

We assumed that infected individuals could be diagnosed from the *Asymptomatic*, *Symptomatic*, or *Severe* states, and that diagnosis would not affect disease progression. We assumed that diagnosis in the Asymptomatic state only occurs among individuals who will not progress to the Symptomatic state. The daily number of these diagnoses is denoted *A_t_* (with the ^ used to indicate quantities related to diagnosis). The fraction of these individuals diagnosed (*q_A,t_*) was assumed to vary over time, to allow for changes in case ascertainment over the course of the epidemic. The delay to diagnosis was defined by , , which is described by a Gamma distribution.

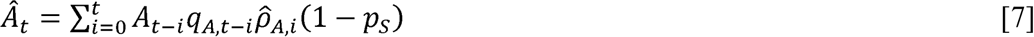

To estimate the number diagnosed from the *Symptomatic* state (*S_t_*) we assumed a time-varying probability of diagnosis , and delay to diagnosis , .

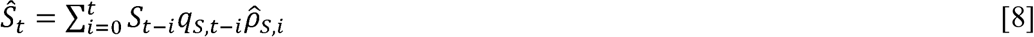

The number diagnosed from the *Severe* state (*V_t_*), was calculated based on a time-invariant probability of diagnosis ( ) and delay to diagnosis , . These were applied after subtracting individuals developing severe disease who had been previously diagnosed at *Symptomatic* (*V*).

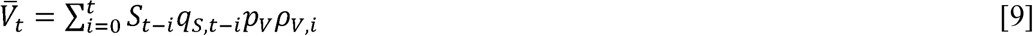

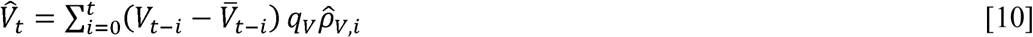

Time-varying diagnosis probabilities (*q_A,t_*, *q_S,t_*, ) were calculated as a function of :

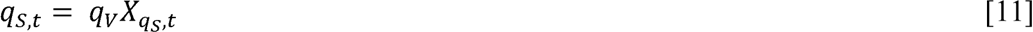

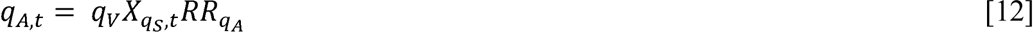

In equations 11 and 12 X*_qs,t_* is a logit-transformed cubic b-spline with knots spaced 21 days apart, with penalties on first and second differences of the spline parameters.*RR_qA_* is constrained to fall in the unit interval, so that that *q_At_* ≤ *q_st_* ≤ *q_v_*for all *t*.

### Reporting

We assumed that all diagnosed COVID-19 cases were reported. The number of diagnoses reported on a given day (C*_t_*, with the ‘·’ used to indicate quantities related to reporting) was calculated as the sum of diagnoses from *Asymptomatic*, *Symptomatic* and *Severe* states, with reporting delay *p_c,i_*

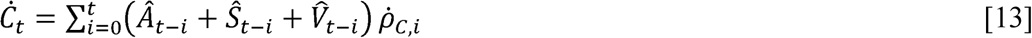

The reported number of COVID-19 deaths (*D_t_*) were calculated from the number of diagnosed individuals who subsequently died (*D_t_* ). *D_t_* was calculated as the sum of deaths among individuals diagnosed from the *Symptomatic* and *Severe* states, represented by the first and second terms in equation 14, respectively. We assumed that all deaths among diagnosed COVID- 19 cases were reported, with reporting delay *p_D,i_*

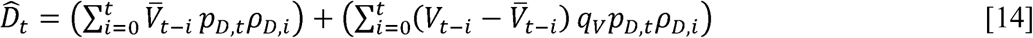

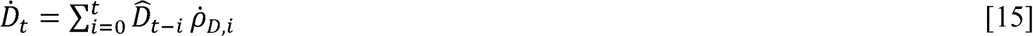

### Data likelihood

We specified negative binomial likelihood functions to fit the model to observed cases (Y*c,t*) and death data (Y*D,t*).

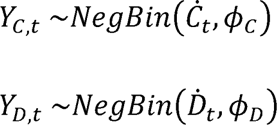

To account for variation in daily reported cases and deaths, we implemented the likelihood using a seven-day moving average of input data and modeled values. The negative binomial dispersion parameters (*ϕ_c_*, *ϕ_D_*) were estimated simultaneously, allowing for additional variance in the observed time series.

### Model parameters

Model parameters are shown in Table 1.

**Table 1:** Model parameters

| Model Parameter | Mean, std. Deviation | Distribution | Type | Source |
| --- | --- | --- | --- | --- |
| Log Of New Infections At T=0 ($A_0$) | 0,10 | Normal(0,10) | prior | Assumed |
| $X_{R,t}$ Spline Parameters | 0,3 | Normal(0,3) | prior | Assumed |
| First Derivative of $X_{R,t}$ Spline Parameters | 0,0.5 | Normal(0,0.5) | prior | Assumed |
| Second Derivative of $X_{R,t}$ Spline Parameters | 0,0.1 | Normal(0,0.1) | prior | Assumed |
| Serial Interval | 5.8, 0.5 | Gamma(129.1, 22.25) | prior | 29 |
| Probability of Developing Symptoms If Infected | 0.59, 0.16 | Beta(5.14, 3.53) | prior | 30-33 |
| Probability of Becoming Severely Ill If Symptomatic | 0.09, 0.06 | Beta(1.89, 20.00) | prior | 35 |
| Probability of Death for All Infections (national average) | 0.005, 0.001 | Beta(15.9, 3167) | prior | 26,27 |
| Probability of Death for Severe Infections | 0.15, 0.03 | Beta(28.2, 162.3) | prior | 35 |
| Rate Ratio, Diagnosis at Asymptomatic Vs. Symptomatic | 0.1, 0.07 | Beta(2,18) | prior | Assumed |
| Rate Ratio, Diagnosis at Symptomatic Vs. Severe | 0.5, 0.22 | Beta(2,2) | prior | Assumed |
| Probability of Diagnosis at Severe | 0.72, 0.16 | Beta(20,5) | prior | Assumed |
| Case Dispersion Parameter $(1/\sigma)^2 *$ | 0.8, 0.6 | Half-Normal(0,1) | prior | 36 |
| Death Dispersion Parameter $(1/\sigma)^2 *$ | 0.8, 0.6 | Half-Normal(0,1) | prior | 36 |
| Time from Infected to Symptomatic (Days) | 5.6, 3.1 | Gamma(3.41, 0.61) | fixed | 37 |
| Time from Symptomatic to Severe (Days) | 7.5, 5.8 | Gamma(1.72, 0.22) | fixed | 38 |
| Time from Severe to Death (Days) | 9.1, 6.3 | Gamma(2.10, 0.23) | fixed | 39 |
| Scaling Factor: Time to Diagnosis Relative to Time in Symptomatic State | 0.5, 0.22 | Beta(2,2) | prior | Assumed |
| Scaling Factor: Time to Diagnosis Relative to Time in Severe State | 0.5, 0.22 | Beta(2,2) | prior | Assumed |
| Case Reporting Delay | 2.2, 1.5 | Gamma(2.2, 1) | prior | Assumed |
| Death Reporting Delay | 2.2, 1.5 | Gamma(2.2, 1) | prior | Assumed |

### Model implementation

The model was implemented in R using the rstan package.^40^ The model initializes 28 days before the first reported case or death. Given the delay from infection to death, we chose 28 days to allow the model to generate the necessary number of new infections to plausibly result in a death early in the observed time series. The model is fit to data from each county or state separately.

For state-level results we estimated outcomes using a Hamiltonian Monte Carlo algorithm.^41^ The model ran for 3000 iterations (2000 burn-in) on 5 chains, and 4000 samples (across 4 chains) from the posterior were included in these results. Counties were fit using an optimization routine that reports the maximum a posteriori estimate, which represents an estimate of the mode of the posterior distribution of the model parameters.

### covidestim Package

The *covidestim* package is a package for the R programming language, suitable for public as well as research use. It can accommodate a number of data inputs. Users may enter a vector of daily case counts and/or daily death counts. These data sources can be used in combination, so long as they are the same length and cover the same time period; days with no observed events may be represented with zeroes.

The package contains default model priors for progression probabilities and delays, detection probabilities and delays, and reporting delays associated with each data type. Users have the ability to override these defaults, though we recommend that they only specify priors for reporting delays; we do not recommend that users change default priors on parameters related to the natural history of COVID-19.

### Covidestim.org and code repositories

We produce daily estimates of COVID-19 infections and the effective reproduction number of SARS-CoV-2 at the state- and county-levels at https://covidestim.org. To allow for daily production of model estimates for all U.S. counties and states, we developed several tools. The *covidestim* Docker image is a container which allows for model execution in any HPC or cloud environment, and is the easiest way to begin using the *covidestim* R package. The *covidestim- sources* repository enables automated, version-controlled, reproducible data cleaning of four different case/death data sources by leveraging Git’s submodules feature. Finally, the *dailyFlow* repository uses the Nextflow workflow engine^42^ to clean the data, orchestrate 3200+ model runs within three supported execution environments (local, HPC, cloud), and export the results for research use and for web consumption. These repositories can be found at https://github.com/covidestim, and contain extensive documentation.

## Data Availability

All data used in the main analysis are available from The Covid Tracking Project and Johns Hopkins CSSE.

https://covidtracking.com/

https://www.mass.gov/info-details/covid-19-response-reporting

https://github.com/covidestim/covidestim

## Acknowledgments

We thank Jeffrey Eaton for his thoughts on statistical analysis.

## Author contributions

NAM and TC conceived and supervised the project. NAM and JS acquired funding. MR, KG, and JH curated data. MHC, MR, FK, VEP, JS, JW, DW, TC, and NAM contributed to the development of the methodology. MHC, MR, FK, and NAM wrote the software. MHC, MR, and NAM conducted the formal analysis. MHC, MR, and NS visualized results. Validation was performed by NAM and NS. MHC and NAM drafted the original manuscript. All authors contributed to the review and editing of the original manuscript.

## Data and materials availability

All data used in the main analysis are available for use at https://github.com/covidestim/covidestim-sources. The *covidestim* package is available for download at https://github.com/covidestim/covidestim.

## References

1. Coronavirus in the U.S.: Latest Map and Case Count. Retrieved from https://www.nytimes.com/interactive/2020/us/coronavirus-us-cases.html

2. Coronavirus. Retrieved from https://www.washingtonpost.com/graphics/2020/national/coronavirus-us-cases-deaths/?itid=hp_pandemic%20test

3. Oran DP and Topol EJ. Prevalence of asymptomatic SARS-CoV-2 infection: a narrative review. Annals of internal medicine. 2020; 173(5): 362–367.

4. Hitchings MDT, Dean NE, Garcia-Carreras B, Hladish TJ, Huang AT, Yang B, Cummings DAT. The Usefulness Of SARS-CoV-2 Test-Positive Proportion As A Surveillance Tool. American Journal of Epidemiology. 2021; 190(7):1396–1405. https://doi.org/10.1093/aje/kwab023

5. Pitzer VE, Chitwood MH, Havumaki J, Menzies NA, Perniciaro S, Warren JL, et al. The impact of changes in diagnostic testing practices on estimates of COVID-19 transmission in the United States. American Journal of Epidemiology. 2021; https://doi.org/10.1093/aje/kwab089

6. Weinberger D, Cohen T, Crawford F, Mostashari F, Olson D, Pitzer VE, et al., Estimating the early death toll of COVID-19 in the United States. [Preprint]. 2020 [Cited 2020 July 13] Available from: https://doi.org/10.1101/2020.04.15.20066431.

7. Gostic KM, McGough L, Baskerville E, Abbott S, Joshi K, Tedijanto C, et al. Practical considerations for measuring the effective reproductive number, Rt. PLoS Comput. Biol. 2021; 16(12): e1008409. https://doi.org/10.1371/journal.pcbi.1008409

8. Unwin HJT, Mishra S, Bradley VC, Gandy A, Mellan TA, Coupland H, et al. State-level tracking of COVID-19 in the United States. Nat. Commun. 2020; 11, 6189 https://doi.org/10.1038/s41467-020-19652-6

9. COVID-19 Portal, Center for the Ecology of Infection Diseases, University of Georgia [Cited 2021 July 10]. Available at: https://www.covid19.uga.edu/nowcast.html

10. Considerations for implementing and adjusting public health and social measures in the context of COVID-19. Geneva: World Health Organization; https://apps.who.int/iris/bitstream/handle/10665/336374/WHO-2019-nCoV-Adjusting_PH_measures-2020.2-eng.pdf?sequence=1&isAllowed=y

11. Ibarrondo FJ, Fulcher JA, Goodman-Meza D, Elliot J, Hofmann C, Hausner MA, et al. Rapid Decay of Anti-SARS-CoV-2 Antibodies in Persons with Mild Covid-19. N Engl J Med. 2020; 383:1085–1087. DOI: 10.1056/NEJMc2025179

12. Seow J, Graham C, Merrick B, Acors S, Pickering S, Steel KJA. Longitudinal observation and decline of neutralizing antibody responses in the three months following SARS-CoV-2 infection in humans. Nat Microbiol. 2020; 5:1598–1607 https://doi.org/10.1038/s41564-020-00813-8

13. Bajema KL, Wiegan RE, Cuffe K, Patel SV, Iachan R, Lim T. Estimated SARS-CoV-2 Seroprevalence in the US as of September 2020. JAMA Intern Med. November 24, 2020; doi:10.1001/jamainternmed.2020.7976

14. Dong E, Du H, Gardner L. An interactive web-based dashboard to track COVID-19 in real time. Lancet Inf. Dis. May 1, 2020; 20(5):533-534. DOI: https://doi.org/10.1016/S1473-3099(20)30120-1

15. Nationwide Commercial Laboratory Seroprevalence Survey. [Cited 23 March, 2020] Available at: https://data.cdc.gov/Laboratory-Surveillance/Nationwide-Commercial-Laboratory-Seroprevalence-Su/d2tw-32xv

16. Wajnberg A, Amanat F, Firpo A, Altman DR, Bailey MJ, Mansour M. Robust neutralizing antibodies to SARS-CoV-2 infection persist for months. Science. 2020; 370(6521):1227–1230. doi: 10.1126/science.abd7728.

17. Qureshi AI, Baskett WI, Huang W, Lobanova I, Naqvi SH, Shyu C. Reinfection with Severe Acute Respiratory Syndrome Coronavirus 2 (SARS-CoV-2) in Patients Undergoing Serial Laboratory Testing. Clinical Infectious Diseases. 2021; https://doi.org/10.1093/cid/ciab345

18. Li T, White LF. Bayesian back-calculation and nowcasting for line list data during the COVID-19 pandemic. PLOS Computational Biology. 2021; 17(7): e1009210. https://doi.org/10.1371/journal.pcbi.1009210

19. Flaxman S, Mishra S, Gandy A, Unwin JT, Mellan TA, Coupland H, et al. Estimating the effects of non-pharmaceutical interventions on COVID-19 in Europe. Nature. 2020; 584: 257–261 https://doi.org/10.1038/s41586-020-2405-7

20. Leung K, Wu JT & Leung GM. Real-time tracking and prediction of COVID-19 infection using digital proxies of population mobility and mixing. Nat Commun. 2021; 12: 1501 https://doi.org/10.1038/s41467-021-21776-2

21. Kishore N, Taylor AR, Jacob PE, Vembar N, Cohen T, Buckee CO, et al. The relationship between human mobility measures and SAR-CoV-2 transmission varies by epidemic phase and urbanicity: results from the United States. [Preprint] 2021. [Cited 15 July 2021]. Available from: https://www.medrxiv.org/content/10.1101/2021.04.15.21255562v1

22. Nouvellet P, Bhatia S, Cori A, Ainslie KEC, Baguelin M, Bhatt S, et al. Reduction in mobility and COVID-19 transmission. Nat Commun. 2021; 12: 1090 https://doi.org/10.1038/s41467-021-21358-2

23. COVID Data Tracker. “Variant Proportions” [Cited 8 June 2021] Available at: https://covid.cdc.gov/covid-data-tracker/#variant-proportions

24. The COVID Tracking Project. [Cited 15 January 2021] Available at: https://covidtracking.com/

25. National Center for Health Statistics. “Provisional COVID-19 Death Counts by Sex, Age, and State” [Cited 15 January 2021] Available at: https://data.cdc.gov/NCHS/Provisional-COVID-19-Death-Counts-by-Sex-Age-and-S/9bhg-hcku

26. O’Driscoll M, Ribeiro Dos Santos G, Wang L, Cummings DAT, Azman AS, Paireau J, et al. Age-specific mortality and immunity patterns of SARS-CoV-2. Nature. 2021; 590; 140–145 https://doi.org/10.1038/s41586-020-2918-0

27. Levin AT, Hanage WP, Owusu-Boaitey N, Cochran KB, Walsh SP, Meyerowitz-Katz G. Assessing the age specificity of infection fatality rates for COVID-19: systematic review, meta-analysis, and public policy implications. Eur J Epidemiol. 2020; 35: 1123–1138. https://doi.org/10.1007/s10654-020-00698-1

28. Razzaghi H, Wang Y, Lu H, et al. Estimated County-Level Prevalence of Selected Underlying Medical Conditions Associated with Increased Risk for Severe COVID-19 Illness — United States, 2018. MMWR Morb Mortal Wkly Rep 2020;69:945–950. DOI: http://dx.doi.org/10.15585/mmwr.mm6929a1

29. X He, EH Lau, P Wu, Marshall KE, Dowling NF, Paz-Bailey G, et al. Temporal dynamics in viral shedding and transmissibility of COVID-19. Nature medicine. 2020; 26(5): pp.672–675.

30. Poletti, P., Tirani, M., Cereda, D., Trentini, F., Guzzetta, G., Sabatino, G., Marziano, V., Castrofino, A., Grosso, F., Del Castillo, G. and Piccarreta, R., 2020. Probability of symptoms and critical disease after SARS-CoV-2 infection. arXiv preprint arXiv:2006.08471.

31. Oran, D.P. and Topol, E.J., 2020. Prevalence of asymptomatic SARS-CoV-2 infection: a narrative review. Annals of internal medicine, 173(5), pp.362–367.

32. Buitrago-Garcia D, Egli-Gany D, Counotte MJ, Hossmann S, Imeri H, Ipekci AM, et al. Occurrence and transmission potential of asymptomatic and presymptomatic SARS-CoV-2 infections: A living systematic review and meta-analysis. PLOS Medicine. 2020; 17(9): e1003346. https://doi.org/10.1371/journal.pmed.1003346

33. O Byambasuren, M Cardona, K Bell, J Clark, M McLaws, P Glasziou (2020). “Estimating the extent of asymptomatic COVID-19 and its potential for community transmission: systematic review and meta-analysis.” JAMMI. 5(4): 223–234. https://doi.org/10.3138/jammi-2020-0030

34. Verity R, Okell LC, Dorigatti I, Winskill P, Whittaker C, Imai N,et al. Estimates of the severity of coronavirus disease 2019: a model-based analysis. The Lancet Infectious Diseases. 2020; 20(6): 669 – 677.

35. CDC COVID-19 Response Team. “Severe Outcomes Among Patients with Coronavirus Disease 2019 (COVID-19) — United States, February 12–March 16, 2020.” MMWR. Morbidity and Mortality Weekly Report (2020) 69(12), 343--346. ISSN 0149-2195, 1545- 861X, doi: 10.15585/mmwr.mm6912e2

36. Gelman, A. Prior Choice Recommendations. Github. 2020 April 17 [Cited 2021 July 10]. Available from: https://github.com/stan-dev/stan/wiki/Prior-Choice-Recommendations

37. SA Lauer, KH Grantz, Q Bi, et al. “The Incubation Period of Coronavirus Disease 2019 (COVID-19) From Publicly Reported Confirmed Cases: Estimation and Application.” Annals of Internal Medicine (2020). 172(9), 577--582. ISSN 0003-4819, 1539-3704, doi: 10.7326/M20-0504

38. F Zhou, T Yu, R Du, et al. “Clinical course and risk factors for mortality of adult inpatients with COVID-19 in Wuhan, China: a retrospective cohort study.” The Lancet. (2020). 395(10229), 1054--1062. ISSN 01406736, doi: 10.1016/S0140-6736(20)30566-3

39. NM Linton, T Kobayashi, Y Yang, K Hayashi, AR Akhmetzhanov, S Jung, B Yuan, R Kinoshita, H Nishiura. “Incubation Period and Other Epidemiological Characteristics of 2019 Novel Coronavirus Infections with Right Truncation: A Statistical Analysis of Publicly Available Case Data.” Journal of Clinical Medicine. (2020). 9(2), 538. ISSN 2077-0383, doi: 10.3390/jcm9020538

40. Stan Development Team. RStan: the R interface to Stan. 2018 R package version 2.17.3. http://mc-stan.org

41. Hoffman MD and Gelman A. The No-U-Turn sampler: adaptively setting path lengths in Hamiltonian Monte Carlo. J Mach Learn Res. 2012; 15(1): 1593–1623.

42. Di Tommaso P, Chatzou M, Floden E, Barja PP, Palumbo E, Noterdame C. Nextflow enables reproducible computational workflows. Nat Biotechnol. 2017; 35, 316–319. https://doi.org/10.1038/nbt.3820

