## Supplementary material for "Reconstructing the course of the COVID-19 epidemic over 2020 for US states and counties: results of a Bayesian evidence synthesis model": Figure S1

Supplementary Materials

Figure S1: Scatterplot of estimates of the infected population plotted as a function of the infection fatality rate, using North Dakota as an example*.

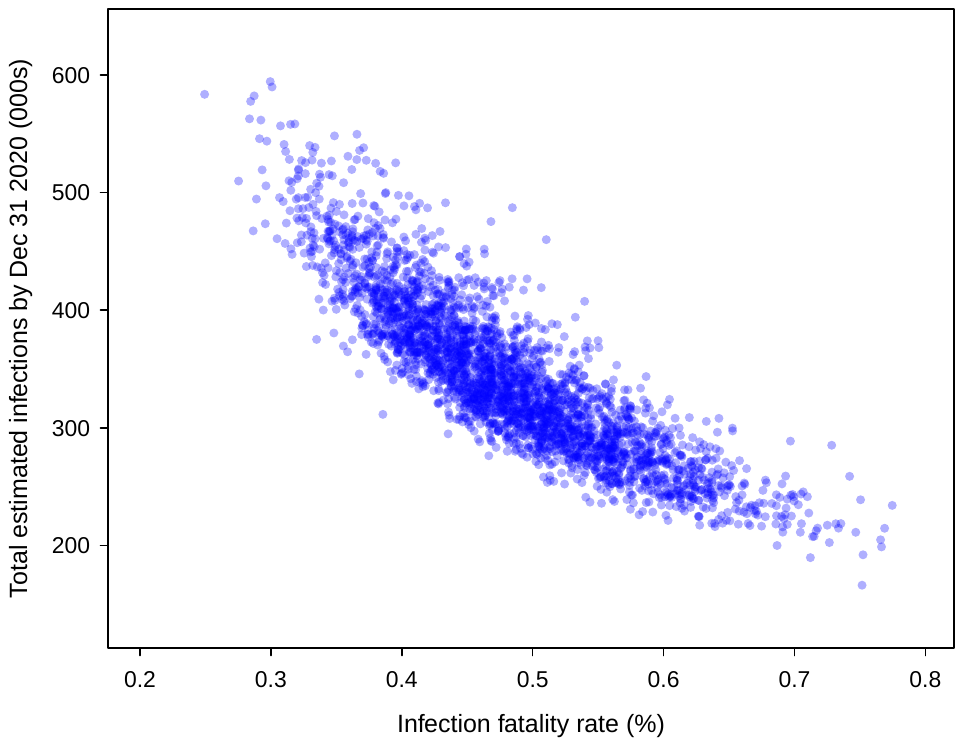

* Each point represents a single draw from the sample of parameter sets generated by the model fitting procedure. Rank correlation = 0.89.
